# Are we really targeting and stimulating DLPFC by placing transcranial electrical stimulation (tES) electrodes over F3/F4?

**DOI:** 10.1101/2022.12.01.22282886

**Authors:** Ghazaleh Soleimani, Rayus Kuplicki, Jazmin Camchong, Alexander Opitz, Martin P Paulus, Kelvin O Lim, Hamed Ekhtiari

## Abstract

**Background:** In many clinical trials involving transcranial electrical stimulation (tES), target electrodes are typically placed over DLPFC with the assumption that this will primarily stimulate the underlying brain region. However, our study aimed to evaluate the electric fields (EF) that are actually delivered and identify prefrontal regions that may be inadvertently targeted in DLPFC tES.

**Methods:** Head models were generated from Human Connectome Project database’s T1+T2-weighted MRIs of 80 healthy adults. Two common DLPFC montages were simulated; symmetric-F4/F3, and asymmetric-F4/Fp1. Averaged EF was extracted from (1) the center of target electrode (F4), and (2) the top 1% of voxels showing the strongest EF in individualized EF maps. Inter-individual variabilities were quantified with standard deviation of EF peak location/value. Similar steps were repeated with 66 participants with methamphetamine use disorder (MUDs) as an independent clinical population.

**Results:** In healthy adults, the group-level location of EF peaks was situated in the medial-frontopolar, and the individualized EF peaks were positioned in a cube with a volume of 29cm3/46cm3 (symmetric/asymmetric montages). EFs in frontopolar area were significantly higher than EF “under” the target electrode in both symmetric (peak:0.41±0.06, F4:0.22±0.04) and asymmetric (peak:0.38±0.04, F4:0.2±0.04) montages (Heges’g>0.7). Similar results with slight between-group differences were found in MUDs.

**Conclusions:** We highlighted that in common DLPFC tES montages, in addition to inter-individual/inter-group variability, the frontopolar received the highest EFs rather than DLPFC as the main target. We specifically recommended considering the potential involvement of the frontopolar area as a mechanism underlying the effectiveness of DLPFC tES protocols.

**Highlights:**

- There are many published tES studies assuming targeting DLPFC with DLPFC tES montages
- In DLPFC tES montages, electric field peaks are not located under the electrodes
- The electric field peak in DLPFC montages is located in the medial frontopolar area
- An independent population showed between-group similarities and variations in electric field
- There is a large interindividual variation in both location and strength of the electric field peak

## 1. Introduction

DLPFC is a commonly used target in non-invasive brain stimulation methods including transcranial magnetic (TMS) or electrical (tES) stimulation (as known as tDCS with low intensity direct current stimulation) [1], [2]. While TMS studies have developed personalized approaches for DLPFC targeting [3], using neuro-navigation systems [4] or fMRI-informed target selection [5], tES studies still widely rely on the 10-20 EEG system, using F4/F3 scalp coordinates to target the right/left DLPFC [6], [7]. Most clinical trials place tES electrodes over the scalp with the assumption that the target electrode (e.g., F4/F3) will dominantly stimulate the underlying brain region (e.g., DLPFC). Consequently, the effects of placing electrodes over F4/F3 have been commonly attributed to modulation of the right/left DLPFC, even in recent publications [8], [9] and less attention has been paid to the possibility that a strong EF in other regions could contribute to the observed clinical/behavioral outcomes of DLPFC-targeting protocols.

Since 2009, gyri-precise head models have demonstrated that tES electrodes produce diffuse current flow, resulting in maximal electric fields (EFs) falling outside the target electrodes rather than directly underneath them [10]–[12]. For example, for tDCS over the primary motor cortex in a single subject simulation, peak EF was not observed directly underneath the large electrode pads, but at an intermediate lobe [10]. Similarly, another single-subject simulation study, which employed atlas-based parcellation, showed that maximum values of tangential and normal components of EFs in DLPFC stimulation were situated in the orbital and frontopolar cortices when electrodes were placed over F3-Fp2 [13]. In group-level analyses, Csifcsak et al also found that in commonly used bipolar DLPFC montages for depression, strong EFs were not only present in DLPFC but also in the medial prefrontal cortex (MPFC) [14], which has been suggested as a new target for depression in TMS studies [15], [16]. In addition, a network-led approach indicated that, although DLPFC is a component of the executive control network, the limbic network received the highest EFs in both symmetric and asymmetric DLPFC tES montages [17].

Additionally, prior research on group-level modeling has also highlighted the significance of inter-individual variability [14], [18], [19]. Considerable differences in morphological features such as skull thickness, cortex morphology, and gyrification have been associated with inter-individual variability [20], [21]). Antonenko et al demonstrated that in DLPFC stimulation EFs differed between young and older adults with higher variability in the former group, and overall EF strength was related to head, skull, and skin volume [19]. Between-group variations in prefrontal stimulation were also observed between clinical and non-clinical populations and people with major depressive disorders and schizophrenia showed lower prefrontal EFs compared to healthy controls [22]. The non-restriction of tES-induced EFs to the area under the electrode and variations in EF distribution patterns may account for disparities in treatment outcomes. Hence, a more comprehensive understanding of the distribution and strength of the peak EFs as a highly stimulated brain region in a large sample size could assist in the development of more effective protocols.

In this context, the primary goal of this study is to evaluate the site/strength of the maximum tES-induced EFs in the prefrontal cortex at a group-level while examining two commonly used electrode montages for DLPFC stimulation (anode/cathode over F4/F3 and F4/Fp1) in both clinical and healthy populations. This work (1) estimates the EF distribution patterns in targeted and non-targeted brain areas in tES over DLPFC, (2) highlights the importance of taking into account tES-induced EFs on brain regions not situated beneath the stimulating electrodes; specifically frontopolar in DLPFC stimulation, and (3) supports previous model-driven approaches for tES target identification.

## 2. Materials and Method

### 2.1. Participants

Unprocessed T1 and T2-weighted structural MRIs from 80 (44 female) randomly selected healthy adults were obtained from the freely available Human Connectome Project database (HCP, http://www.humanconnectomeproject.org/data/) with deidentified anatomical scans; age (year) between 31-35 (n = 32, 15 female), between 26-30 (n = 34, 21 female), between 22-25 (n = 14, 8 female) (the exact age of participants are not reported in the database). Additional details on MRI parameters are available in the supplementary materials.S1.

### 2.2. Creation of head models and EF simulations

High-resolution T1 and T2-weighted MRIs were combined to create individualized computational head models for all 80 participants using the standard SimNIBS 3.2 pipeline and the “headreco” function [23]. However, three participants were excluded from the study due to encountering an error during the mesh generation process. Gmsh failed to mesh one or more surfaces in 3 attempts for two participants. In another participant, mesh generation failed in decoupling between white matter and gray matter ventricles. Consequently, all results were reported based on EFs for 77 healthy participants. DLPFC montages were simulated by placing 5×7 cm electrodes with 1 mm thickness over (1) F4/F3 (symmetric montage) and (2) F4/Fp1 (asymmetric montage) locations as two frequently used DLPFC montages. Further details on generating head models and EF calculations can be found in the supplementary materials.S2.

### 2.3. Data analysis

For each individual, averaged EF strength was extracted from the main regions of interest (ROIs) in the prefrontal cortex, as outlined in section 2.4. Peak EF was defined as the 99^th^ percentile of the EF over the cortex and both location and strength of the peak were extracted at both individual and group levels. Between-subject variations were quantified using the relative standard deviation (SD) of the EFs strength and locations. Numerical statistical analyses were performed using the *R* package. Since the Shapiro-Wilk test of normality showed our database is normally distributed, t-tests were used to examine significant differences between EFs in two separate brain regions. All data reported as mean ± SD. Statistical results were presented based on P values and Hedges’ g factors to explain the significance level and effect sizes.

### 2.4. Defining regions of interest (ROI)

In order to compare EF strength over DLPFC and around EF peaks, two individualized 10 mm spheres were defined: (1) around the center of the target electrode (F4) location, and (2) around 99^th^ percentile of the EFs for each individual. Spheres were combined with the MNI mask to ensure analyses did not include EFs from non-brain or white matter voxels. Brainnetome atlas was also used for defining regions of interest in the prefrontal cortex based on atlas-based parcellation of the head models (supplementary materials.S3).

### 2.5. Replication of the results with a sample clinical population

Based on our updated systematic review on transcranial electrical stimulation trials (tES) in substance use disorders (SUDs), by the end of 2022, 72 out of 85 (85%) tES studies in SUDs used bipolar DLPFC stimulation with symmetric or asymmetric montages [24]. Over the years various positive effects have been reported but there is also a decent number of studies that find no effects for the application of tES to reduce drug craving or consumption [25]–[27]. These variations could be due to individual variability and experimental variability. However, less attention has been paid to inter-individual variability in terms of tES-induced EFs in people with SUDs. In order to determine the replicability of the results in a clinical population and in order to identify brain regions that received the highest EFs in people with SUDs, all previous steps were repeated for 66 participants (age (year) between 18-60, all male) with methamphetamine use disorders (MUDs) as a representative example of a clinical population (more details about participants, T1 and T2 MRIs, and data collection protocols can be found in our previously published paper [17]; this database included only male participants and MRI data for women were not available).

## 3. Results

### 3.1. Comparing EFs at the center of target electrodes and hot spots

Individual and group-level analysis of personalized head models showed that the 99^th^ percentile of the EFs (peaks) is not located underneath the stimulating electrodes (F4; [40.5, 41.4, 27] coordinate over the cortex in MNI space). Averaged locations for the peak EFs across the population were [-1.21, 57.66, 18.67] for symmetric and [-2.62, 49.00, -4.54] for asymmetric montages (refer to section 3.2). With respect to the brain anatomical subregions, EF peaks were located within the frontopolar area; a region occupying the anterior portion of the brain’s frontal lobe (dominantly corresponding to Brodmann’s area 10) which is distinct from DLPFC (which is located on the lateral and dorsal part of the medial convexity of the frontal lobe [28]) where the stimulating electrode was placed (dominantly comprises Brodmann’s areas 9 and 46) [29].

EFs were also extracted in the coordinates related to the individualized 99^th^ percentile and center of the stimulating electrode (Figure 1). EF strength in the coordinate related to the 99^th^ percentile (symmetric = 0.41±0.06, asymmetric = 0.38±0.05) was significantly (P value = 2.2×10^-16 for both montages) higher than the coordinate related to the center of stimulating electrode (symmetric = 0.23±0.05, asymmetric = 0.20±0.04) with large effect sizes (Hedges’g for: symmetric = 3.2091 with 95% CI (2.73, 3.69) and asymmetric = 3.8339 with 95% CI (3.30, 4.37)). EFs were also significantly higher in the symmetric montage compared to asymmetric (P value for: 99th percentile = 0.004, the center of target electrode = 1.7×10^-5) with small effect sizes for 99th percentile individualized coordinate (Hedges’g = 0.4724 with 95% CI (0.15, 0.79)) and medium effect size for the coordinate related to the center of target electrode (Hedges’g = 0.7114 with 95% CI (0.34, 1.04)).

**Figure 1.**
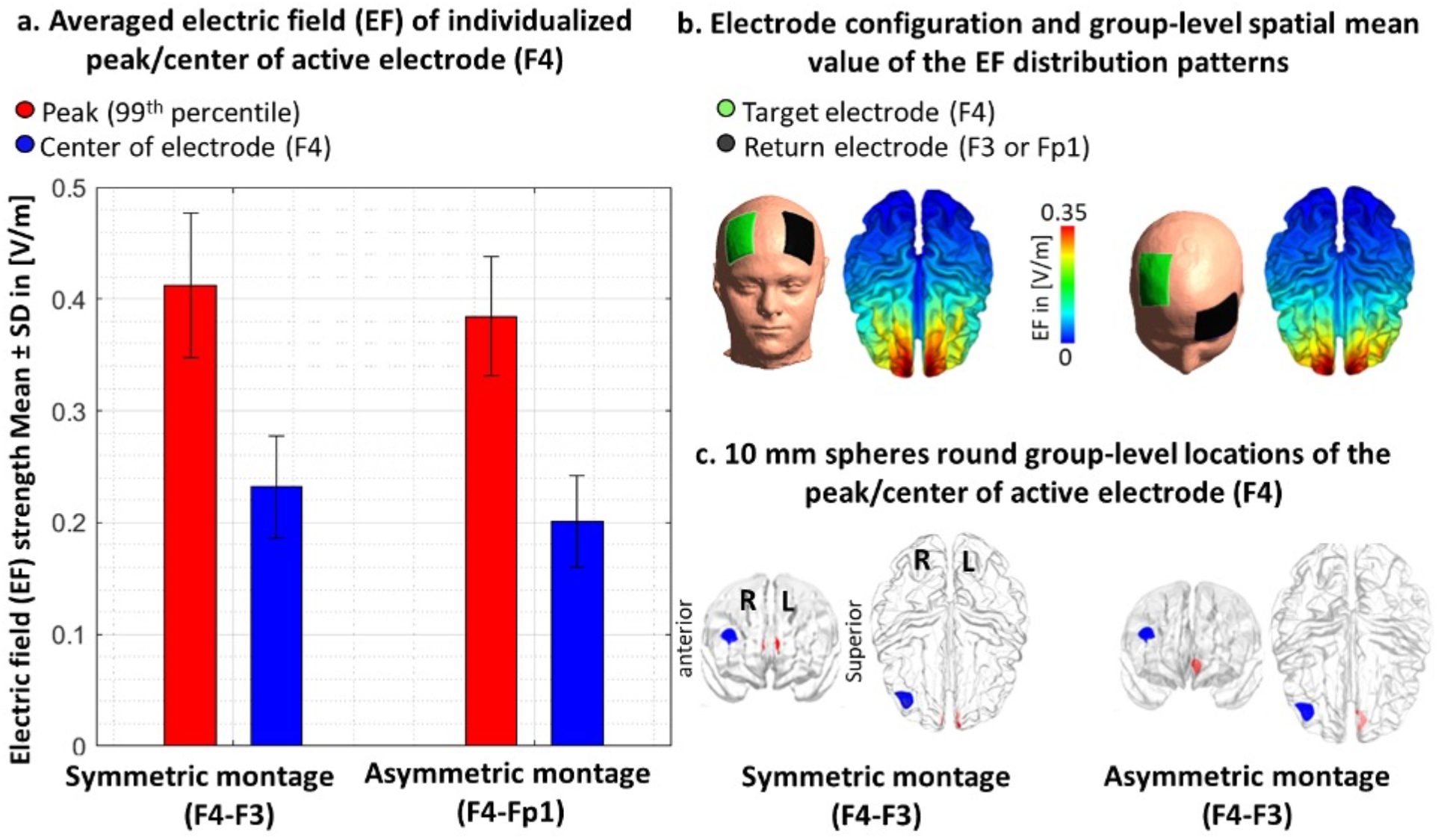
Comparing EF (electric field) in the center of the target electrode vs. peak EF. **(a)** Bars show mean values and error bars show standard deviations (SD) of the EF strength in volts per meter ([V/m]) across 77 healthy adults in individualized 99th percentile of the EF (peak EF, in red) and center of the target electrode (F4, in blue) over the cortex across the population for symmetric (target/return electrodes over F4/F3 in dark colors) and asymmetric (target/return electrodes over F4/Fp1 in light colors) montages. **(b)** Electrode configurations over the scalp for DLPFC stimulation are visualized with the target (in green)/return (in black) electrodes over F4/F3 in symmetric and F4/Fp1 in asymmetric montages. EF distribution patterns at the group-level (spatial mean values across the population) are visualized over the cortex in superior view. **(c)** Locations of the 10 mm spheres around F4 (in blue) and averaged location of the peak EF across the population (in red) are visualized over the standard brain in fsaverage space. In each panel, left side corresponds to the symmetric montage (F4-F3) and the right side corresponds to the asymmetric (F4-Fp1) montage.

Results were also calculated based on averaging EFs in 10mm spheres around the peak and F4 locations and the same results were found in terms of significantly higher EFs in the frontopolar area compared to DLPFC (supplementary materials.S4). In addition to the spherical ROIs, averaged EFs were also extracted from the main regions of the prefrontal cortex using Brainnetome atlas parcellation and cumulative EF strength with a symmetric montage in the right frontopolar was significantly (P<0.01 with Hedges’ g = 0.3412) higher than right DLPFC (supplementary materials.S5, Figure.S1).

### 3.2. Inter-individual variabilities

Inter-individual variability in terms of peaks’ locations and strength was compared across the population. Variations across the population are visualized in Figure 2. In the symmetric DLPFC stimulation, the mean location for the peaks across the population in MNI space was [-1.21, 57.66, 18.67] with [6.43, 4.37, 8.08] as SD, and all peak locations were placed in a cube with a volume of 29 cm3 (X = 3.1, Y = 1.9, Z = 4.9). Group-level mean value for the EF strength was also 0.41±0.06 [V/m] (ranges from 0.31 to 0.68). In the asymmetric DLPFC stimulation, the mean location for the peaks in MNI space was [-2.62, 49.00, -4.54] with [7.05, 7.71, 8.94] as SD, and all peak locations were placed in a cube with a volume of 46 cm3 (X = 3.3, Y = 3.4, Z = 4.1). Group-level mean value for the EF strength was also 0.38±0.04 [V/m] (ranges from 0.28 to 0.59).

**Figure 2.**
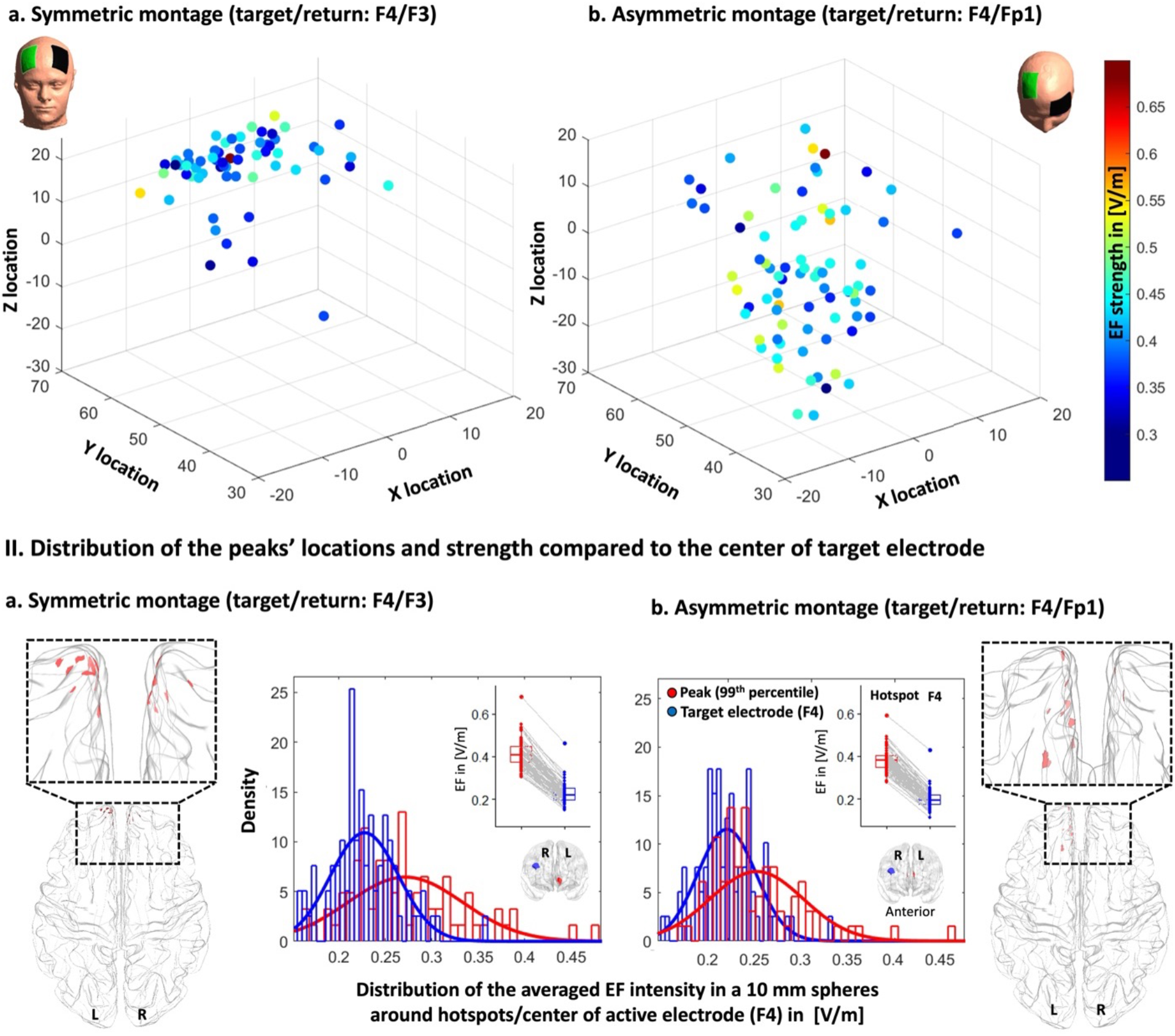
Inter-individual variability of electric field (EF) strength and locations. **I**. Scatter plot (for location in MNI space) colored based on EF strength (hot colors represent strong EF strength) for visualizing inter-individual variability in peaks (99th percentile) of the EFs in (a) symmetric and (b) asymmetric DLPFC montages. **II**. Visualizing the location of the 99th percentile of the EF over the standard fsaverage space (red dots over the cortex represent each individual). Distribution plots represent the distribution of the EF strength within the 10 mm spheres around F4 and 99th percentile (blue and red spheres over the cortex in anterior view) for (a) symmetric and (b) asymmetric montages. Boxplots showing differences between EFs in the coordinates related to individualized peaks and F4 in [V/m]. Dots and spaghetti lines over the boxplots represent the data for individual participants. Abbreviation: EF: electric field.

### 3.4. Replication results in a clinical population

All steps were repeated for the MUD group. In the symmetric montage averaged location for the EF peaks in MNI space was [1.88, 60.34, 19.12] with [5.71, 3.34, 6.72] as SD, and all peaks were placed inside a cube with a volume of 21.7 cm^3^. In the asymmetric montage averaged location of the EF peaks in MNI space was [-3.14, 56.90, 6.33] with [7.37, 5.86, 9.74] as SD, and all peaks were placed in a cube with a volume of 45.9 cm^3^. Our EF strength results (Figure S2) showed that EF strength in the frontopolar area (symmetric: 0.40±0.07, asymmetric: 0.41±0.06) where EF peaks were placed is significantly (P<0.001) higher than DLPFC (symmetric: 0.19±0.05, asymmetric: 0.21±0.05) underneath the target electrode with large effect sizes (Hedges’g for symmetric = 1.6190 with 95% CI (1.19, 2.04) and asymmetric = 1.3599 with 95% CI (0.95, 1.77)). However, no significant difference (P>0.1) was found between the EFs strength in symmetric and asymmetric montages in this population such that effect sizes were negligible (for peak EFs Hedges’g = 0.13 with 95% CI (−0.24, 0.50) and around F4 location Hedges’g = -0.02 with 95% CI (−0.39, 0.34)). Additionally, by considering inter-individual variability (Figure S3), in atlas-based parcellation of the head models, results (Figure S4) showed that the frontopolar area (A10l+A10m) received a significantly higher EF strength compared to DLPFC (A9/46v+A9/46d) in both symmetric and asymmetric montages (P<0.001).

Our results showed that averaged peak location in symmetric (healthy: [-1.21, 57. 66, 18.67], MUDs: [1.88, 60.34, 19.12] with maximum 4.12 mm^3^ Euclidean distance from F4 location over the cortex) and asymmetric montages (healthy: [-2.62, 49, -4.54], MUDs: [-3.14, 56.9, 6.33] with maximum 13.45 mm^3^ Euclidean distance from F4 location over the cortex) have a substantial overlap between the two groups such that in both groups peaks were located within the frontopolar area (Figure 3). Furthermore, using an unpaired t-test for between-group comparison in terms of EF strength in spherical regions around peaks (10mm spheres were defined around the averaged peaks’ locations and F4 in MUD and healthy then mean EFs were extracted from the 10mm sphere), a significantly higher EF strength in the asymmetric montage in the MUD (0.27±0.07) was found compared to the healthy group (0.23±0.05) with a medium effect size (Hedges’ g = 0.6172 with 95% CI (0.27,0.97)). A significant difference in the symmetric montage was also found around the F4 location with a higher EF strength in the healthy group (0.23±0.05) compared to MUDs (0.19±0.04) with a large effect size (Hedges’ g = -0.9059 with 95% CI (−1.27,-0.55)).

**Figure 3.**
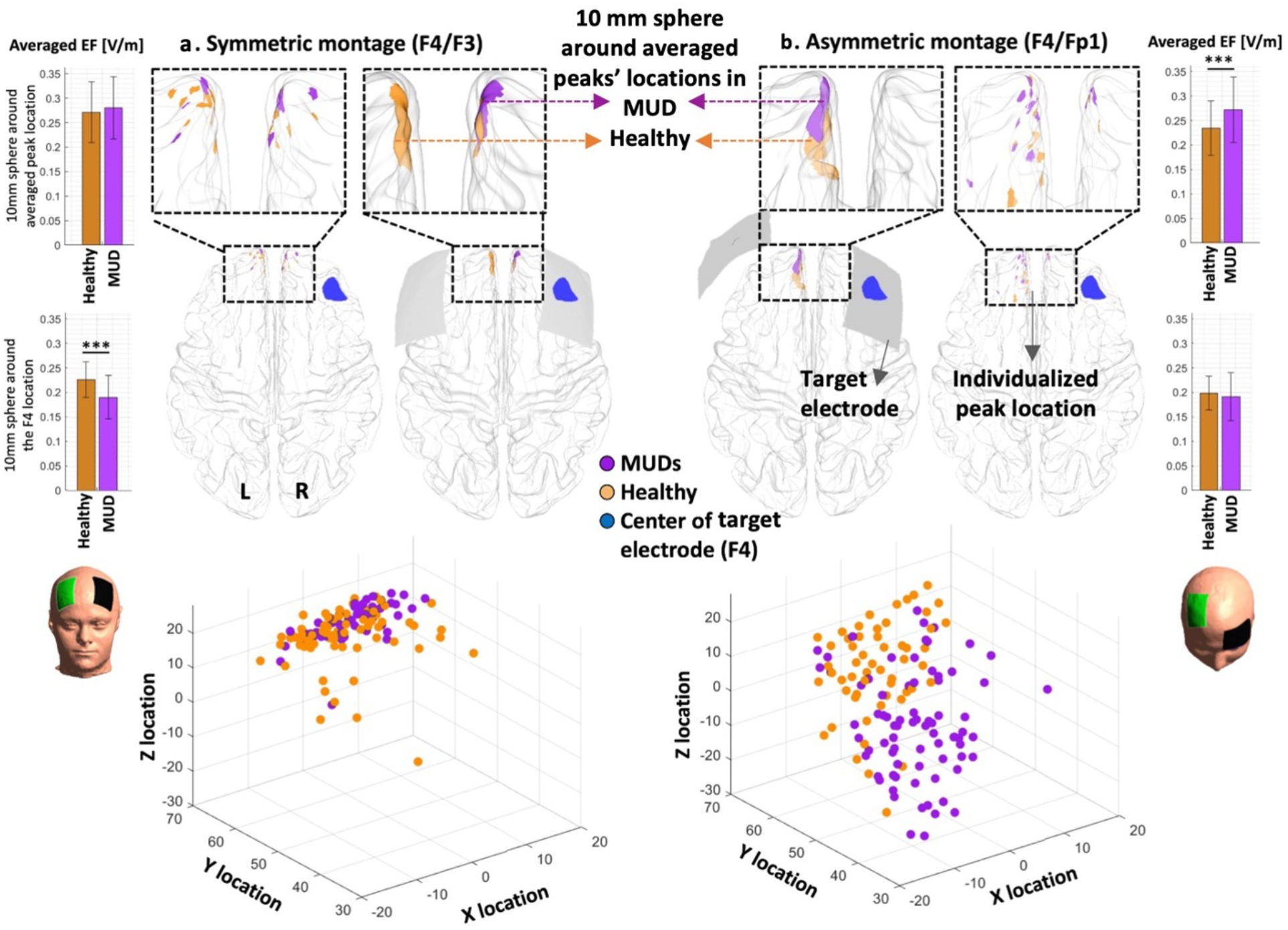
Comparison between healthy participants and methamphetamine users in terms of location and strength of the peaks at both individual and group levels. Overlap between 10 mm spheres around mean EF peaks in healthy participants (in brown) and a group of participants with methamphetamine use disorders (in purple) compared to the center of the target electrode F4 (in blue) in both symmetric (left panel) and asymmetric (right panel) montages. Small dots represent each individual in each group and big circles represent 10 mm spheres around averaged peaks’ locations at the group-level. MNI coordinates for F4 = [40.5, 41.4, 27], averaged peak location in symmetric montage (F4/F3): healthy participants = [-1.21, 57.66, 18.67] and methamphetamine users = [1.88, 60.34, 19.12] with 4.12 mm3 Euclidean distance, asymmetric montage (F4/Fp1): healthy participants = [-2.62, 49, -4.54], methamphetamine users = [-3.14, 56.90, 6.32] with 13.45 mm3 Euclidean distance.

## 4. Discussion

In this study, we investigated the effectiveness of two commonly used DLPFC montages (F4/F3: symmetric, F4/Fp1: asymmetric) for tES in inducing electric fields by analyzing their target selectivity. We aimed to determine whether DLPFC is the main target and to what extent other brain regions that are not intentionally targeted may receive an electric field (EF) above the primary target. Individualized computational head models were generated for two groups of participants; healthy adults and people with methamphetamine use disorders (MUDs). Specifically, this investigation yielded four main results. Firstly, the peaks of EF were located far (45.54 Euclidian distance in MNI space) from DLPFC (F4 location as the center of target electrode). Secondly, group-level EF peaks were dominantly placed near the medial frontopolar cortex in both montages, and both groups with significantly higher EFs compared to DLPFC. Thirdly, variations in peak EF location and strength were observed within and between groups. Finally, our results supported previous model-driven approaches for tES target determination and enabled assessing how EF distribution patterns in DLPFC stimulation in SUDs may affect different parts of the brain including the frontopolar area.

### 4.1. Importance of peak EFs in brain stimulation studies

Our computational approach with a substantial sample size underlies the utility of head models for uncovering potentially stimulated brain regions based on EF peaks. As investigated in previous brain stimulation studies [30], the efficacy of the stimulation protocol may vary depending on the brain region being modulated. However, systematic analysis of EF distribution patterns and finding the association between tES-induced EFs and stimulation outcomes requires a decision about which EF measures (e.g., maximum, mean, median, or a binary thresholded EF map) are appropriate to use. Here, inspired by previously published studies in the field [31], [32], we focused on EF peaks to provide an indicator of the location at which the target activity is most likely to be perturbed [33]. Then, peak EF location and strength in the peak location were compared with the cortical area under the center of target electrodes as the main “intended” anatomical target.

In conventional tES, it has been repeatedly reported that maximal EF is not underneath the electrodes but rather between them and outside the “area under the electrode” [10], [34], [35]. Due to the diffuse current flow, it’s not surprising that we found EF peaks out of DLPFC; similar to previous studies on motor cortex stimulation that reported maximum EFs between two electrodes [36]. However, our data suggest that top 1% of voxels with the highest EF are located farther from the intended cortical target DLPFC and specifically are located in the frontopolar area. Therefore, if the main goal of the stimulation is reaching the brain area beneath the target electrode (F3/F4), it is recommended to define an ROI in the targeted brain region and extract averaged value from non-thresholded EF maps.

### 4.2. Inter-individual variability

Our results showed significant inter-individual variability in the location/strength of the peak EF in directly/indirectly targeted brain areas which are in line with previous studies that emphasize the importance of considering personalized head models [20], [34], [35]. The main issue with the inter-individual variability of EFs over the cortex is that the potential effect of tES is limited by small effect sizes and it would be difficult to detect intervention effects at the group-level [37]–[39]. Different sources of variability including anatomical parameters (e.g., CSF volume) affect EF distribution patterns [18], [40], [41]. The variability of the simulation results indicates that personalized electrode montages should be considered to have similar cortical stimulation doses inside a target area across a population.

### 4.4. DLPFC montages make peak EF in frontopolar areas

Our findings about peak EF within the frontopolar cortex are in line with a previous modeling study that reported strong EF intensity within the medial prefrontal cortex (MPFC) rather than DLPFC in all commonly used bipolar DLPFC electrode montages in depression [14]. This study suggested that symptom improvement in DLPFC tES trials in depression might not necessarily be specifically related to the DLPFC and other brain areas with strong EF (e.g, MPFC) may also contribute to the tES treatment outcomes [14]. In line with this assumption, preliminary evidence from clinical trials confirmed associations between EF strength and behavioral changes in the depression [42]. In a DLPFC stimulation study with electrodes over F5/F6 locations in a group of participants with MDD, a significant positive correlation between EFs within the MPFC and behavioral outcomes was reported while the correlation between EFs in DLPFC and the same behavioral outcome (negative affect) was negative (greater score reductions were associated with lower EF strength) [42]. Taken together, a brain region with higher EFs might be linked to behavioral outcomes through several psychophysiological mechanisms. Therefore, based on our simulations, we argue that conventional electrodes over DLPFC also stimulated the frontopolar area. We recommend taking into account the cognitive process associated with the frontopolar region in interpretation of the results in conventional tES studies over the DLPFC [18], [20], [40], [41].

### 4.6. Different EF patterns in DLPFC montages

Another finding in this study was significant differences between symmetric and asymmetric montages in terms of EF strength in healthy participants. As we discussed in our previous publication with a network-based perspective [17], cathode location significantly affects EFs strength in targeted or non-targeted brain areas [35], [43]. Here, we found that symmetric DLPFC stimulation induced significantly higher EFs in both DLPFC and peaks compared to asymmetric montage. Lower EF strength in asymmetric montages could be related to a lower distance between the electrodes over the scalp compared to the symmetric montage [44]. It can be attributed to more current shunting in asymmetric montage compared to symmetric since electrodes are in closer proximity [45]. More significant changes in behavioral outcomes using symmetric DLPFC stimulation studies might be related to the stronger cortical EFs compared to asymmetric montages. For instance, previous studies showed that symmetric montages over DLPFC could diminish risk-taking behavior during a Balloon Analogue Risk Task (BART) [46], [47], while asymmetric DLPFC stimulation shows no such effect [48]. Between montage difference in terms of peaks’ strength only in the healthy group, not MUDs, suggest that generalization of the results obtained from EF distribution patterns fundamentally depends on the population of interest and highlights the importance of head modeling in tES protocol optimization in both individual and group levels [19], [49].

### 4.7. Atlas-based vs spherical regions of interest

Our study shows that the results obtained from defining regions of interest (ROI) based on peak and center of target electrode locations align with the atlas-based parcellation results. Although the most common approach for exploratory ROI analysis (e.g., in fMRI analysis) is to create small spheres around the peaks’ locations (e.g., peaks of activation clusters), atlas-based parcellation may help to simply explore and localize informative regions (e.g., subregions of the prefrontal cortex involved in DLPFC stimulation) based on the spatial distribution patterns of the EFs. Our analysis of EFs over multiple nodes in the prefrontal cortex using atlas-based parcellation revealed the potential for synergistic stimulation outcomes through excitatory/inhibitory pathways between anatomically distinct brain regions. However, as reported by [50], the specific brain parcellation of each atlas impacts the spatial accuracy of the extracted brain regions especially when differences in the observed EF distribution patterns are small. To ensure accuracy, inspiered by [22], we used the Brainnetome atlas, which provides a fine-grained parcellation of the prefrontal cortex. Nonetheless, the regions specified by atlas-based parcellation can be large (e.g., SFG in Brainnetome atlas), and even if highly modulated nodes are present, strong EFs may occur only in a small proportion of voxels within the ROI. Therefore, averaging across a parcel may lose spatial detail about the EF. Personalized EF distribution patterns or using a fine-grained atlas, such as the Schaefer atlas [51], maybe a better approach for defining ROIs compared to averaging through a large parcellated brain area.

### 4.7. Optimized targeting: evidence from HD-tES, neuroimaging, and TMS studies

Here, we only focused on large electrode pads while using focal electrode montages could improve the targeting of the stimulation site [10]. However, a previous systematic review reported a lack of evidence for the effectiveness of focal tES stimulation, for example with high-definition (HD) electrodes, in currently published clinical trials [52]. While HD electrodes may enhance the spatial focality [10], achieving more focal EF comes at the cost of lower intensity and increased inter-individual variability [35]. Factors such as electrode position, electrode size/shape/orientation, and between-electrode distance can affect EF distribution patterns and focality [35]. Rather than relying solely on fixed HD montages, optimization algorithms could be used to identify a multi-array electrode arrangement that maximally targets a predefined region while minimizing EFs in nontargeted areas [53].

Using neuroimaging data such as fMRI that help to understand the causal role of targeted and non-targeted brain regions [54], is also a promising approach toward more efficient and precise target selection [55], [56]. This approach has gained momentum recently, particularly in the field of TMS, and could potentially offer valuable information about functional localization to optimally guide EF peak locations based on modulating currently active neural circuits [57], [58]. Closing the loop between brain-state and stimulation parameters could also enhance the efficiency of the optimization using concurrent tES-fMRI. In this context, stimulation dose (e.g., stimulation intensity, frequency, and phase difference) could be optimized at an individual level to maximally stimulate the ongoing brain state [59].

### 4.9. SUDs as a sample clinical population

Results of simulation for the MUD group indicated the presence of differences between groups, which could be attributed to alterations in brain anatomy observed in people with SUDs compared to healthy participants [49], [60]. Our results are in line with previous between-group differences in clinical and non-clinical populations and suggest that EF distribution patterns cannot be simply transferred from healthy participants to people with MUDs [22]. However, similar to healthy participants, in both DLPFC montages, frontopolar received the highest EFs in the MUD group. This suggests the frontopolar area may play a role in tES over DLPFC for SUDs while much of the initial attention is focused on the DLPFC with more promising results with symmetric compared to asymmetric montages [24]. The importance of targeting the frontopolar area in SUDs is further supported by tES studies integrated with fMRI while people with SUDs were exposed to drug cues [61]. Reported results by Nakamura et al demonstrated that the application of bilateral DLPFC increased functional activity in the ventromedial prefrontal cortex (VMPFC not DLPFC) during drug cue exposure [61]. Maybe the reason behind the significant rise in functional activity within VMPFC upon DLPFC stimulation is linked to the highest EF intensity in this particular brain area, which remains unexplored.

Dose-response relationship analysis, which integrated head models with fMRI (refer to supplementary materials.S7), found further support for the importance of EF strength in frontopolar area involvement in DLPFC stimulation for SUDs [50], [62]. A pre-post tDCS-fMRI study on MUD participants showed a significant correlation between changes in brain functions in response to drug cues and the averaged EF strength solely in the frontopolar area, not other prefrontal regions [50]. A larger cohort of individuals with MUDs also showed similar results; a significant correlation was observed between the normal component of the EF and changes in functional activity in the frontopolar area during a standard drug cue reactivity task [62].

The importance of the frontopolar area in brain stimulation for SUDs is also supported by lesion-based studies that reported the frontopolar cortex as a key region in the trans-diagnostically relevant neural circuits contributing to the addiction [63]. Lesion-based fMRI data revealed that a higher likelihood of decreasing substance use was associated with brain injury to areas that had negative connectivity to the frontopolar areas and recommended frontopolar as an ideal neuromodulation treatment target for SUDs [63].

### 4.10. Limitations and future directions

Our study has some limitations that need to be addressed in future research. Firstly, we only employed structural MRI and EF simulations and did not measure other behavioral/neural outcomes to determine the impact of EFs in each targeted and non-targeted brain region. Furthermore, we did not examine the relationship between EFs and clinical responses, such as drug consumption/craving in our clinical population. Open questions remain about how to relate EFs with behavioral or neurophysiological changes in both healthy and clinical populations. Furthermore, since we did not consider age/sex-matched clinical population and because the imaging protocols were different between the two groups, we did not directly compare our results between MUDs and healthy participants as conducted by [22]. Since EF distribution patterns may differ in the field of addiction compared to healthy participants due to the brain anatomical alterations [49], [60], and because variations in skull thickness between sexes could lead to differences in peak EF location/value, comparing EF peaks between SUDs and healthy controls in age and sex-matched case-control cohorts could be investigated in future studies. For instance, our results showed that the center of averaged EF peak location is deeper in the brain of our clinical population compared to healthy participants. The main reason for this outcome could be investigated in future studies.

## 5. Conclusion

Here, we discussed that, in two of the most frequently used electrode montages in tES studies which are commonly intended for modulating DLPFC, DLPFC was not maximally targeted by placing large electrodes over this region. Instead, other parts of the prefrontal cortex received stronger electric field intensity, e.g., the frontopolar cortex. Considering inter-individual variability, our results highlighted the crucial role of the frontopolar area in tES over DLPFC. Based on individual and group-level analysis of EF peaks, in both healthy participants and MUDs, we recommend that future trials should not solely attribute F3/F4 tES outcomes (with symmetric or asymmetric configuration) to the DLPFC. Other brain regions in medial prefrontal areas like medial frontopolar should be considered in upcoming trials placing large electrode pads over DLPFC.

## Data Availability

The raw database and head models generated for this study are available on request to the corresponding author.

## 6. Acknowledgements

This study is supported by funds from Laureate Institute for Brain Research, Tulsa, OK, Medical Discovery Team on Addiction (MDTA), University of Minnesota, Minneapolis, MN and Brain and Behavior Foundation (NARSAD Young Investigator Award #27305) to HE. The grant RF1MH117428 was provided to AO. There was no role for the funding agency in the design, execution, analysis or reporting this study.

## Supplementary materials

### S1. Participants: MRI data

Structural MRI data in the Human Connectome Project (HCP) data archive were collected with a Siemens MAGNETOM 3T scanner with 32 channel head coil and the following parameters for T1-weighted MRIs: TR/TE = 2400/2.14, flip angle = 8, the field of view = 224 × 224 × 180 mm3 and voxel size = 0.7 mm3, and T2-weighted MRIs: TR/TE = 3200/565. More details on imaging parameters and data acquisition can be found in the HCP database, Appendix I: Structural Session Scan Protocol.

### S2. Creation of head models and EF simulations

SimNIBS 3.2 pipeline was used for generating computational head models [1]. Briefly, T1 and T2-weighted images were segmented into six tissue types, including white matter (WM), gray matter (GM), cerebrospinal fluid (CSF), skull, scalp, and eyeballs, using an automated tissue segmentation approach in SPM 12 based on the “headreco” function. Segmentation results were evaluated carefully slice by slice to ensure proper tissue classification. Tetrahedral volume meshes with about 3×106 elements were created for each head model and visualized using Gmsh and MATLAB.

A standard EEG cap (EEG10-10-UI-Jurak-2007) was used for placing electrodes over the scalp. In the symmetric montage, centers of the target electrodes were located over F4 and F3 locations with the long axis of the pads pointing towards the vertex of the head. In the asymmetric montage, the forehead electrode (Fp1) was positioned over the left eyebrow with the long axis of the pad parallel to the horizontal plane (as shown in Figure 1, panel b). Here we focused on 35 cm^2^ rectangular electrodes since this size/shape was by far the most commonly used in previously published studies (e.g., in our updated systematic review in the field of addiction medicine, among total of 67 published studies, only 4 studies used circular while 38 studies used two 5×7 electrode [2]). However, the orientation of non-circular electrodes can affect the strength and direction (normal component) of the EFs over the cortex. Therefore, the locations and orientations of the electrodes were precisely checked in Gmsh to guarantee consistency after automatic placement using a modified MATLAB code in SimNIBS.

Through the SimNIBS pipeline, the finite element method (FEM) was used to simulate EFs for head meshes. Linear and isotropic electrical conductivities were assumed based on previously established conductivity values for each tissue type [3]: white matter = 0.126, gray matter = 0.275, cerebrospinal fluid

= 1.654, skull = 0.010, skin = 0.465, and eyeballs = 0.5 all in Siemens per meter (S/m). The absolute and normal components of the EFs were calculated for each montage and each individual. Surface-based head models were then transformed from individualized space to the “fsaverage” standard space (http://surfer.nmr.mgh.harvard.edu) to make the results comparable across the population in terms of brain coordinates. Although normalization to the standard space can affect EF distribution patterns, our previous study showed no statistically significant difference between the group-level results obtained from standard space and the subject space [4].

### S3. Defining region of interest (ROI): Atlas-based parcellation

The Brainnetome atlas (which has a fine-grained parcellation of the cortex as a multimodal parcellation atlas based on structural MRI, diffusion tensor imaging, and resting-state fMRI connectivity) was used for the parcellation of the computational head models [5]. Inspired by [6], ROIs were placed in 9 main subregions in the Brainnetome atlas; Superior frontal gyrus: A9l, lateral area [13,48,40], A9m, medial area [6,38,35], A10m: medial area [8,58,13]. Middle frontal sgyrus: A9/46d, dorsal area [30,37,36], A9/46v, ventral area [42,44,14], A46 [28,55,17], A10l, lateral area [25,61,-4], and Orbital gyrus: A11l, lateral area [23,36,-18], A11m, medial area [6,57,-16]. Averaged EF strength was calculated for each of the 9 prefrontal subregions. Differences between subregions were calculated using ANOVA and posthoc pairwise t-test.

### S4. Comparing EFs at the center of target electrodes and hot spots: Averaging EFs in 10mm spheres

In both electrode montages, mean EFs in a 10mm sphere around the individualized peak EFs (symmetric: 0.32±0.07, asymmetric: 0.24±0.05; mean ± SD in V/m) was significantly (P value for: symmetric = 3.01×10^-4, asymmetric: 4.52×10^-6) higher than a 10mm sphere around F4 location over the cortex in which the center of the target electrode was placed (symmetric: 0.22±0.05, asymmetric: 0.19±0.05 in V/m) with large effect sizes (Hedges’g for: symmetric = 0.8598 with 95% CI (0.53,1.19) and asymmetric = 0.7630 with 95% CI (0.43,1.09)). Our results also showed that, by averaging EFs in 10 mm spheres around individualized peak EF locations across the population, symmetric montage produced significantly (P = 0.00015) higher EFs compared to asymmetric electrode arrangement with medium effect sizes (Hedges’g for: peak’s location = 0.6234 with 95% CI (0.95, 0.30) and under the target electrode = 0.7965 with 95% CI (0.47, 1.13)).

### S5. Comparing EFs at the center of target electrodes and hot spots: Atlas-based parcellation results

In addition to the spherical ROIs around the center of target electrode (DLPFC) and peaks (medial frontopolar), averaged EFs were also extracted from the main regions of the prefrontal cortex including superior frontal gyrus (SFG), middle frontal gyrus (MFG), and orbital gyrus (OrG) in the Brainnetome atlas (areas related to the Brodmann area 9, 10, 11, and 46 as defined in the method section); SFG: A9l, A9m, A10m, MFG: A9/46d, A9/46v, A46, A10l, OrG: A11m, and A11l. Averaged EFs in each subregion in both left and right hemispheres along with the representation of the regions over the cortex can be found in the supplementary materials (Figure S1). Cumulative EF strength with a symmetric montage in the right frontopolar (A10m + A10l) was significantly (P<0.01 with Hedges’ g = 0.3412) higher than right DLPFC (A9/46v+A9/46d). Additionally, symmetric montage induced slightly higher EFs compared to the asymmetric montage in almost all subregions but differences were significant only in the A10m in both hemispheres (P<0.001).

**Figure S1.**
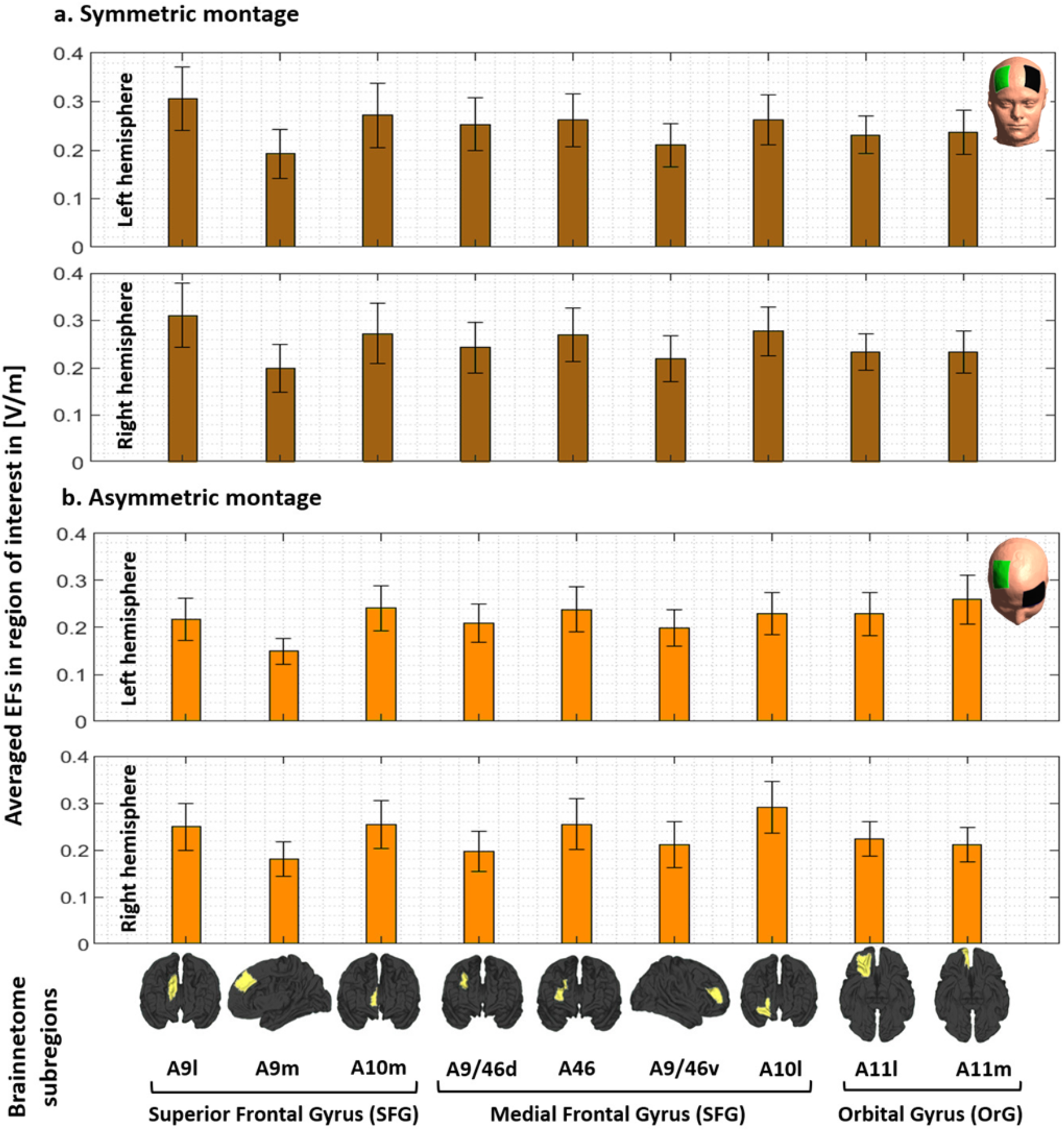
EF strength within the main subregions of the prefrontal cortex. Computational head models were generated for all participants and a Brainnetome atlas with 105 subregions in each hemisphere was used to extract tES-induced EFs in each cortical areas based on **(a)** symmetric (dark brown) and **(b)** asymmetric (light brown) montages. Bars show mean values and error bars show standard deviations (SD) of the EF strength in volt per meter ([V/m]) across 77 healthy subjects in 9 main subregions of the prefrontal cortex in the left (lines 1 and 3) and right (line 2 and 4) hemispheres. Labels below the horizontal axis determine the name of each subregion based on Brainnetome atlas parcellations and small brains next to the labels represent each region in fsaverage space. Atlas-based parcellation: *<u>Superior frontal gyrus:</u>* A9l, lateral area [13,48,40], A9m, medial area [6,38,35], A10m: medial area [8,58,13]. *<u>Middle frontal gyrus:</u>* A9/46d, dorsal area [30,37,36], A9/46v, ventral area [42,44,14], A46 [28,55,17], A10l, lateral area [25,61,-4]. *<u>Orbital gyrus</u>*: A11l, lateral area [23,36,-18], and A11m, medial area [6,57,-16]. Abbreviation: EF: electric field, SFG: superior frontal gyrus, MFG: medial frontal gyrus, OrG: orbital gurus.

### S6. Replication results in a clinical population

All results are presented for the MUD group.

**Figure S2.**
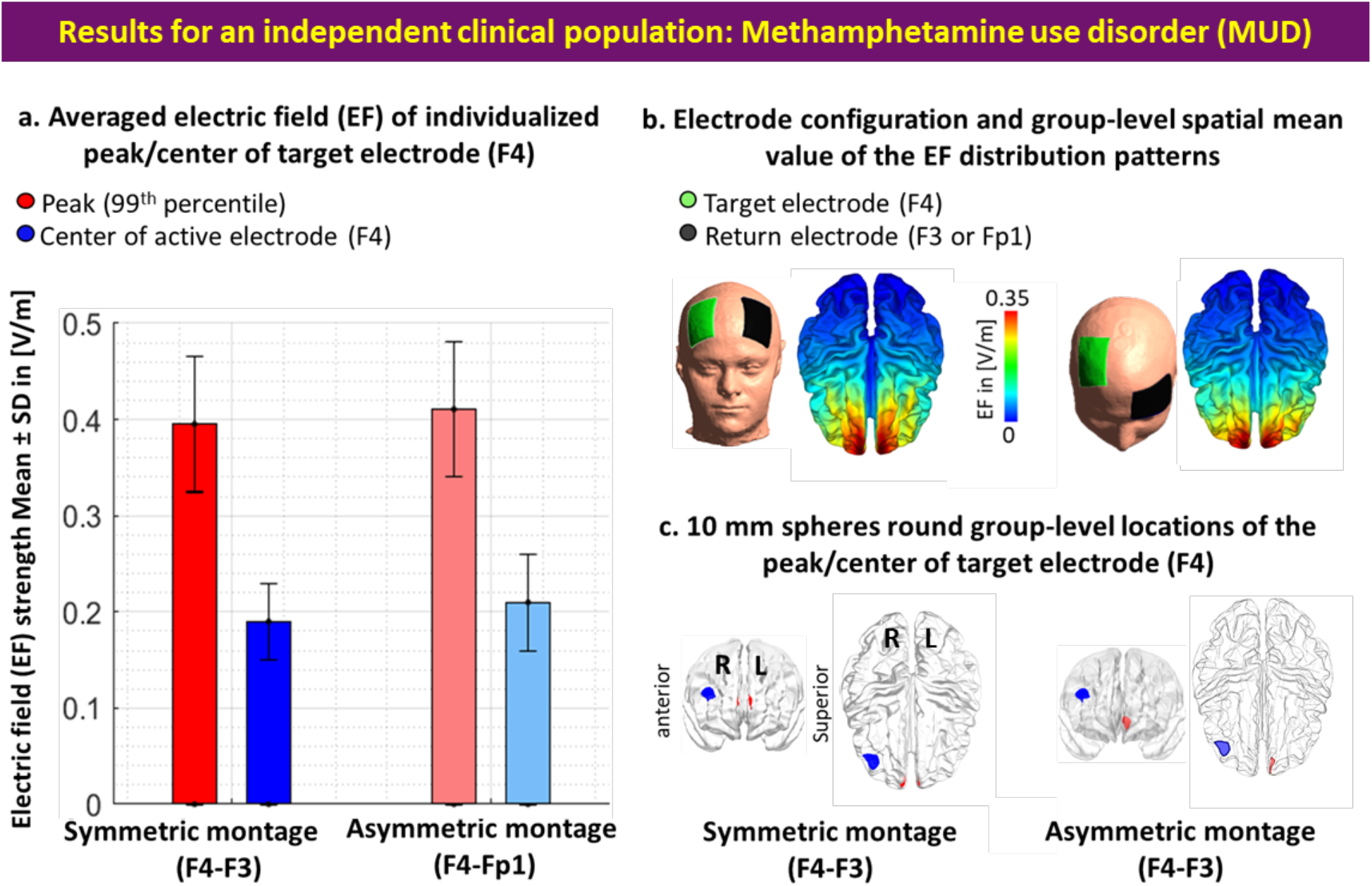
Comparing the center of the target electrode with EF peaks in an independent clinical population-people with methamphetamine use disorder (MUD). **a**. Bars show mean values and error bars show standard deviations (SD) of the EF strength in volt per meter ([V/m]) across 66 participants with MUD in individualized 99^th^ percentile of the EF (peaks, in red) and center of the target electrode (F4, in blue) over the cortex across the population for symmetric (target/return over F4/F3 in dark colors) and asymmetric (target/return over F4/Fp1 in light colors) montages. Target electrodes over F4 are depicted in green, return electrodes over F3 in symmetric, and Fp1 in asymmetric montages are depicted in black. **b**. Electrode configurations over the scalp for DLPFC stimulation are visualized with the target (in green)/return (in black) electrodes over F4/F3 in symmetric and F4/Fp1 in asymmetric montages. EF distribution patterns at the group-level (spatial mean values across the population) are visualized over the cortex in superior view. **c**. Locations of the 10 mm spheres around F4 (in blue) and averaged location of the peaks across the population (in red) are visualized over the standard brain in fsaverage space. In each panel, the left side corresponds to symmetric montage (F4-F3) and right side corresponds to asymmetric (F4-Fp1) montages. Abbreviation: EF: electric field.

**Figure S3.**
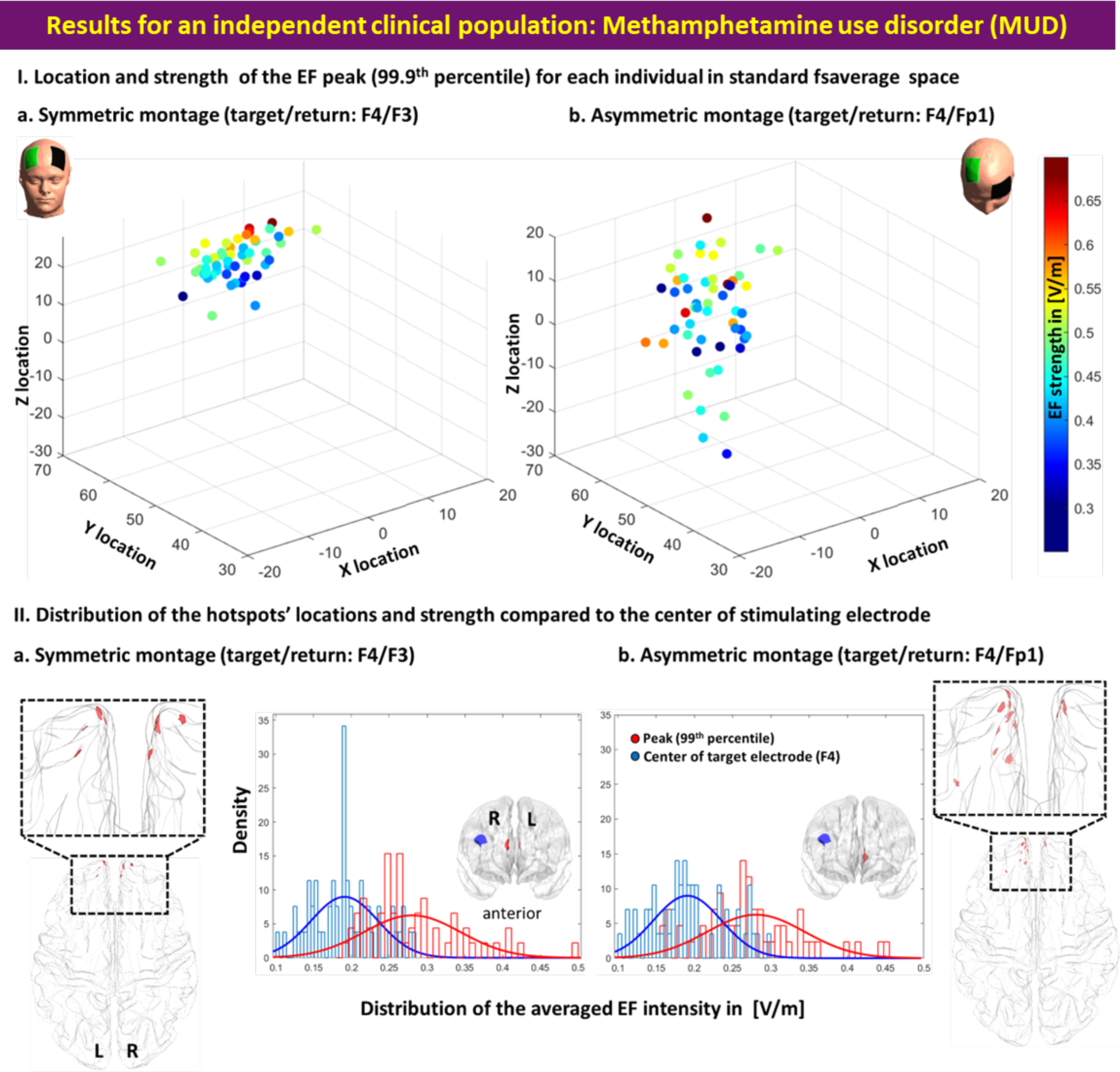
Inter-individual variability of peak EF location and strength in an independent clinical population-people with methamphetamine use disorder (MUD). **I**. Scatter plot (for location in MNI space) colored based on EF strength (hot colors represent strong EF strength) for visualizing inter-individual variability in the 99^th^ percentile of the EFs in symmetric **(a)** and asymmetric **(b)** DLPFC montages. **II**. Visualizing the location of the 99^th^ percentile of the EF over the standard fsaverage space (red dots over the cortex represent each individual). Distribution plots represent the distribution of the EF strength within the 10 mm spheres around F4 and 99^th^ percentile (blue and red spheres over the cortex in anterior view) for symmetric **(a)** and asymmetric **(b)** montages. Abbreviation: EF: electric field.

**Figure S4.**
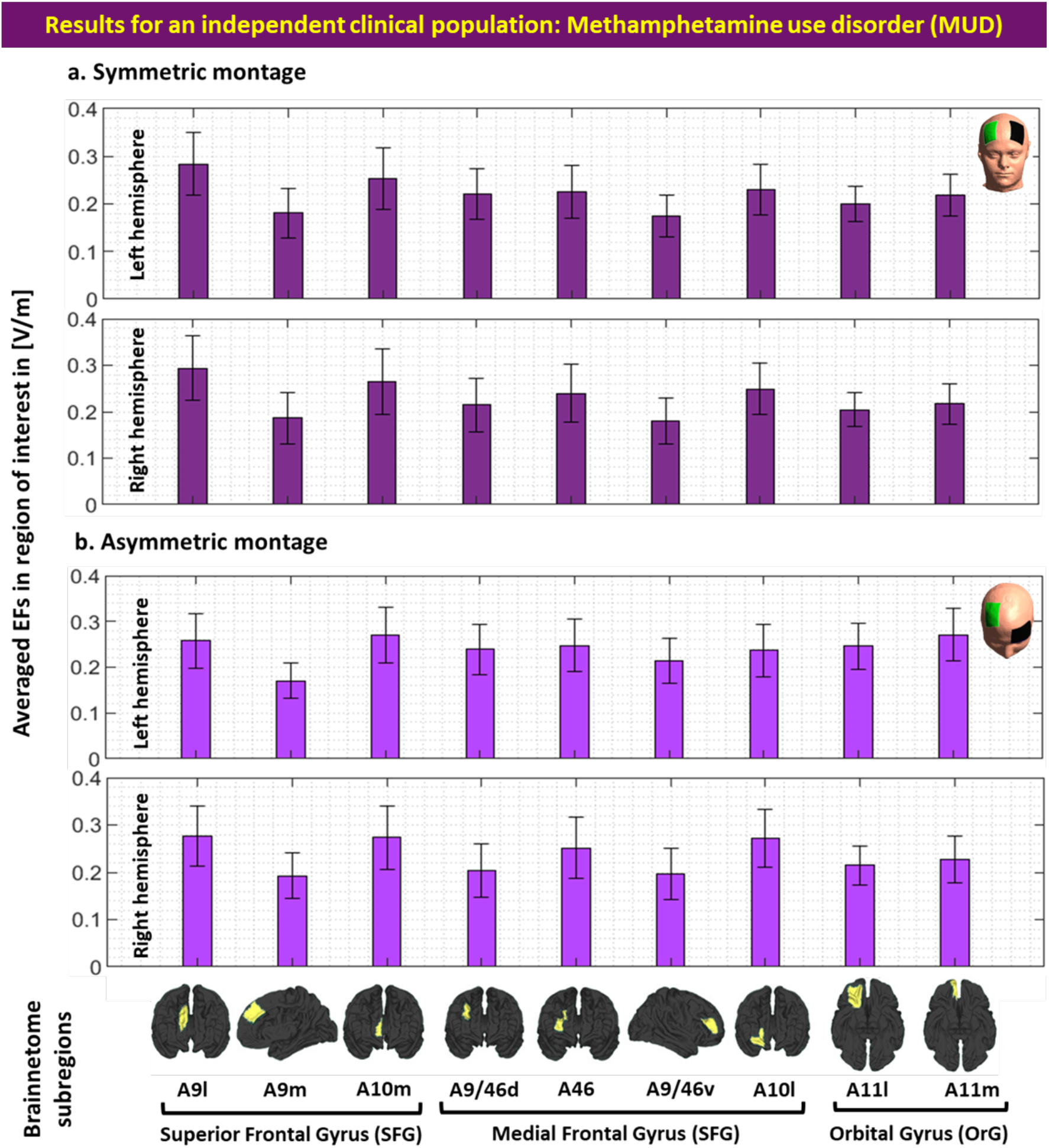
EF strength within the main subregions of the prefrontal cortex in an independent clinical population-people with methamphetamine use disorder (MUD). Computational head models were generated for all participants and a Brainnetome atlas with 105 subregions in each hemisphere was used to extract tES-induced EFs in each cortical areas based on **(a)** symmetric (dark green) and **(b)** asymmetric (light green) montages. Bars show mean values and error bars show standard deviations (SD) of the EF strength in volt per meter ([V/m]) across 66 participants with methamphetamine use disorder (MUD) in 9 main subregions of the prefrontal cortex in the left (line 1 and 3) and right (line 2 and 4) hemispheres. Labels below the horizontal axis determine the name of each subregion based on Brainnetome atlas parcellations and small brains next to the labels represent each region in fsaverage space. Atlas-based parcellation: *<u>Superior frontal gyrus:</u>* A9l, lateral area [13,48,40], A9m, medial area [6,38,35], A10m: medial area [8,58,13]. *<u>Middle frontal gyrus:</u>* A9/46d, dorsal area [30,37,36], A9/46v, ventral area [42,44,14], A46 [28,55,17], A10l, lateral area [25,61,-4]. *<u>Orbital gyrus</u>*: A11l, lateral area [23,36,-18], and A11m, medial area [6,57,-16]. Abbreviation: EF: electric field, SFG: superior frontal gyrus, MFG: medial frontal gyrus, OrG: orbital gurus.

### S7. Dose-response relationship

Here, we only focused on highly modulated brain regions with respect to the strong EF over the cortex using high-resolution structural MRI data. It has been assumed that brain regions with higher EFs contribute to greater alteration in brain responses and may have stronger effects on stimulation outcomes (e.g., higher EFs followed by greater functional alterations) [7]. Under the assumption that EF strength over the cortex relates to the tES responses at the functional level, the association between stimulation dose and brain responses in a cortical target could explain dose-response relationships in tES studies. Recent advancements in neuroimaging (like fMRI) and neurophysiology (like TMS) suggest a complicated non-linear and state-dependent dose-response relationship in tES studies. However, there is still some supporting evidence that highlights the role of brain regions with strong EFs in physiological/neural response to tES that may help to determine the role of DLPFC and frontopolar area and the importance of brain regions with higher EFs in symmetric/asymmetric DLPFC stimulation studies.

One of the earliest studies in which both EFs and fMRI data were discussed was by Halko et al 2011. The single-subject case study of tDCS with combined visual rehabilitation training after stroke revealed that EF strength was correlated with task-based fMRI activation in some predefined ROIs; higher EF was linearly related to stronger functional activation [8]. In a group of participants with left-sided glioma, averaged EF strength was extracted from the left and right M1 ROIs, and a significant correlation between averaged EFs in the right M1 and changes in global resting-state connectivity from the right M1 was reported [9]. Antonenko et al. also assessed the relationship between tES-induced EFs and neurophysiological outcomes and significant negative/positive correlations were reported between the tangential/normal component of the EF and resting-state functional connectivity in the sensorimotor cortex [10]. Additionally, Jamil et al investigated whether cerebral blood flow (CBF) activations across the cortex agree respectively with EFs obtained from head models. Using a voxel-wise rank correlation, a significant correlation between averaged EF distribution patterns in MNI space and the group-level T-contrast images was reported at the voxel-level in both cathodal and anodal stimulation [11]. Recently it has also been shown that current density in the left DLPFC positively correlated with changes in functional connectivity between two predefined ROIs (left dorsolateral and left ventrolateral PFC) during a working memory task based on using psychophysiological interaction analysis and simulating precise computational head models [12]. All previous dose-response relationships highlight the importance of EF strength and its effects on stimulation outcomes that shed light on how modulated brain areas with stronger EFs may affect the response to stimulation. A similar investigation approach (which has not been investigated in healthy subjects) is needed to assess dose-response relationships in the frontopolar area while DLPFC is targeted. In addition to correlational methods, testing for the causal role of the frontopolar in DLPFC stimulation studies can help to confirm the importance of the frontopolar cortex in the regulation of stimulation outcomes.

